# Real-world data on immune responses following heterologous prime-boost COVID-19 vaccination schedule with Pfizer and AstraZeneca vaccines in England

**DOI:** 10.1101/2021.12.14.21267562

**Authors:** Samantha J Westrop, Heather J Whitaker, Annabel A Powell, Linda Power, Corinne Whillock, Helen Campbell, Ruth Simmons, Lenesha Warrener, Mary E Ramsay, Shamez N Ladhani, Kevin E Brown, Gayatri Amirthalingam

## Abstract

**Background:** There are limited data on immune responses to heterologous COVID-19 immunisation schedules, especially following an extended ≥12-week interval between doses.

**Methods:** SARS-CoV-2 infection-naïve and previously-infected adults receiving ChAd-BNT (ChAdOx1 nCoV-19, AstraZeneca followed by BNT162b2, Pfizer-BioNTech) or BNT-ChAd as part of the UK national immunisation programme provided blood samples at 30 days and 12 weeks after their second dose. Geometric mean concentrations (GMC) of anti-SARS-CoV-2 spike (S-antibody) and nucleoprotein (N-antibody) IgG antibodies and geometric mean ratios (GMR) were compared with a contemporaneous cohort receiving homologous ChAd-ChAd or BNT-BNT.

**Results:** During March-October 2021, 75,827 individuals were identified as having received heterologous vaccination, 9,489 invited to participate, 1,836 responded (19.3%) and 656 were eligible. In previously-uninfected adults, S-antibody GMC at 30 days post-second dose were lowest for ChAd-ChAd (862 [95%CI, 694 – 1069]) and significantly higher for ChAd-BNT (6233 [5522-7035]; GMR 6.29; [5.04-7.85]; p<0.001), BNT-ChAd (4776 [4066-5610]; GMR 4.55 [3.56-5.81]; p<0.001) and BNT-BNT (5377 [4596-6289]; GMR 5.66 [4.49-7.15]; p<0.001). By 12 weeks after dose two, S-antibody GMC had declined in all groups and remained significantly lower for ChAd-ChAd compared to ChAd-BNT (GMR 5.12 [3.79-6.92]; p<0.001), BNT-ChAd (GMR 4.1 [2.96-5.69]; p<0.001) and BNT-BNT (GMR 6.06 [4.32-8.50]; p<0.001). Previously infected adults had higher S-antibody GMC compared to infection-naïve adults at all time-points and with all vaccine schedules.

**Conclusions:** These real-world findings demonstrate heterologous schedules with adenoviral-vector and mRNA vaccines are highly immunogenic and may be recommended after a serious adverse reaction to one vaccine product, or to increase programmatic flexibility where vaccine supplies are constrained.

What is already known?
PubMed was searched with the terms “COVID-19 Vaccine” and “heterologous” to identify publications relating to heterologous immunisation schedules with adenoviral-vector and mRNA vaccines from 01 January 2020 until 30 November 2021. Following early reports of vaccine-induced thrombocytosis and thrombocytopenia (VITT) after the first dose of ChAd (ChAdOx1 nCoV-19), several studies reported significantly higher antibody levels, with robust neutralizing activity and cellular immune responses, in adults receiving a heterologous ChAd-mRNA schedule compared to those receiving ChAd-ChAd. Few studies, however, have compared antibody responses after both heterologous schedules (ChAd-mRNA and mRNA-ChAd) with both homologous schedules (ChAd-ChAd and mRNA-mRNA). One UK study (COMCOV) compared all four ChAd and BNT162b2 Pfizer-BioNTech (BNT; mRNA) combinations given four weeks apart and reported very high antibody and T-cell responses four weeks after the second dose for all four schedules.

What are the new findings?
We used the national immunisation register to identify adults who received a heterologous vaccine schedule as part of the national immunisation programme in England and collected blood samples to measure SARS-CoV-2 antibody responses after vaccination. We found that both heterologous schedules (ChAd-BNT and BNT-ChAd) provided superior antibody responses compared to ChAd-ChAd and similar responses to BNT-BNT at 30 days and 12 weeks after second vaccine dose. ChAd-BNT induced higher antibody levels then BNT-ChAd at both timepoints. Antibody responses after vaccination were much higher in previously infected individuals, irrespective of their immunisation schedule. A recent Swedish population-based study reported higher vaccine effectiveness against symptomatic disease with ChAd-BNT than ChAd-ChAd providing real-world confirmation of improved protection with heterologous schedules.

What do the new findings imply?
Our findings add to the growing body of evidence showing high antibody responses following heterologous vaccination schedules with ChAd and BNT, along with robust antibody neutralising activity and cellular responses, especially when compared to ChAd-ChAd. Given that globally COVID-19 vaccine demand far exceeds vaccine supply, these results have important implications for the future deployment of COVID-19 vaccine programmes; particularly where it is logistically and/or operationally difficult to administer two doses of the same vaccine product.

## Introduction

COVID-19 vaccines are highly effective in preventing severe disease and deaths due to SARS-CoV-2. There are currently more than twenty vaccines that have been approved and rolled out globally (1). The United Kingdom was one of the first countries to implement a national COVID-19 immunisation programme in December 2020, initially with BNT162b2 (BNT, Pfizer BioNTech), a nucleoside modified mRNA vaccine, and soon followed by AstraZeneca ChAdOx1/nCoV-19 (ChAd, AstraZeneca), which utilises a simian adenovirus vector. Pre-licensure clinical trial data demonstrated high humoral and cellular responses after a two-dose schedule with high vaccine effectiveness against symptomatic disease (2) (3). The UK, like most other countries, recommended immunisation with the same vaccine brand for both doses where possible, although a heterologous prime-boost vaccine schedule was advised for a small number of individuals in specific circumstances, such as serious adverse events after the first dose, including anaphylaxis (4). Following rare reports of, vaccine-induced thrombocytosis and thrombocytopenia (VITT) after the first dose of ChAD, many countries recommended completing the schedule with an mRNA vaccine for younger adults who had received an adenoviral vector vaccine for their primary dose (5).

Furthermore, given that most of the global population remains unvaccinated, the option to offer a heterologous schedule could potentially simplify logistics of program delivery; helping to mitigate against supply chain issues, and support populations to increase second dose coverage. There are, however, limited data on comparing different heterologous COVID-19 extended vaccine schedules.

We and others have reported increased reactogenicity rates after the second dose using heterologous compared to homologous vaccine schedules (6) (7), and there are increasing reports of comparable or improved humoral and cellular responses following heterologous schedules with ChAd and BNT when compared to two doses of ChAd (8) (9) (10). Direct comparisons of different mixed extended schedules, however, are limited. In Spain, the CombiVacS trial reported that anti-S protein antibodies were successfully boosted upon administration of a heterologous booster (BNT) 8-12 weeks after a priming dose of ChAd, with an acceptable reactogenicity profile (8). In Germany, those who received heterologous BNT boost 9-12 weeks following vaccination with ChAd demonstrated significantly higher neutralising antibody levels at 14 days post-boost compared to those receiving BNT-BNT or ChAd-ChAd (9). In England, the COMCOV study recruited adults to four study arms to receive one of each of the four possible combinations of ChAd and BNT (10), but these were administered with a 28-day interval, rather than an UK-recommended extended schedule with a 12 week interval between doses (12).

We therefore undertook real-world serological assessment of the immunogenicity of heterologous compared to homologous COVID-19 vaccination schedules in adults receiving different combinations of ChAd and BNT vaccines at 30 days and 12 weeks after the second dose of vaccine.

## Methods

The UK Health Security Agency (UKHSA), formerly Public Health England (PHE), has been conducting national COVID-19 surveillance in England throughout the pandemic. Individuals aged 15-75 years recorded to have a heterologous COVID-19 vaccine schedule between 29 March 2021 and 17 September 2021 were identified through the National Immunisation Management System (NIMS) – a real-time national electronic database containing records of all individuals receiving a COVID-19 vaccine in England which is updated daily. Potential participants were initially recruited in London, the South East and East of England and then extended nationally, as described previously (6) (12). Those with a second vaccine dose recorded in the previous 21 days and a mobile phone number or email address in NIMS were invited to take part by text message/email. A link was shared to provide information about the evaluation. Those willing and eligible to participate were asked to sign an electronic consent form online and complete a short online questionnaire, which was developed using SnapSurvey software.

A pragmatic approach was taken for blood sample collection; including venepuncture by a trained health care professional or use of a self-sampling capillary blood collection device (TASSO-SST/TASSO-PLUS). Participants using self-sampling devices were requested to obtain the blood sample as soon as they received the kit and return the sample by post in UN3373 compliant packaging to the national Virus Reference Department, UKHSA, on the same day. Reminders were sent to those who did not return the kit promptly.

### Inclusion/Exclusion Criteria

Individuals who confirmed receiving two different COVID-19 vaccine products, as evidenced on their COVID-19 immunisation record card, were eligible to take part. Those who were unable to provide informed consent, self-reported as being immunosuppressed due to disease or treatment (4), were unwilling to use a self-sampling device (TASSO-SST) or did not agree to attend for phlebotomy, were excluded. Individuals who reported taking anticoagulants and who were not able to attend for phlebotomy were also excluded because of the unsuitability of the TASSOPLUS device for self-sampling in this circumstance.

### Patient and public involvement

This evaluation was rapidly deployed in response to the SARS-CoV-2 pandemic to inform the UK national immunisation programme. As such no members of the public were involved in the design, analysis or dissemination of results. At any stage participants were able to contact the study team via phone and/or email with any queries, concerns or feedback, and such information was used to improve the study.

### Sample time points

Blood samples were requested at 30 days (±9 days) and 12 weeks (±2 weeks) after the second vaccine dose. If participants were unable to provide an adequate sample at 30 days, they were invited to repeat the sample if still within the sampling window or provide a sample at 12 weeks. Returned samples obtained outside of the sampling windows were not included in the analysis.

### Comparator groups

SARS-CoV-2 antibodies in adults aged 50-74 years who were enrolled in the CONSENSUS study and received homologous vaccination as part of the UK national immunisation programme were used as comparator groups. These data have been published and are included here for comparison only (11).

### Assessment of antibody levels

IgG antibody levels against the SARS-CoV-2 spike protein (S-antibody) and Nucleoprotein (N-antibody) were determined using the Roche Elecsys S and Roche N assays respectively (13) (14). Roche anti-S IgG were expressed as arbitrary units (au)/mL serum with a positive threshold of ≥ 0.8 (13).

### SARS-CoV-2 infection

As part of the online questionnaire, participants were asked to report previous SARS-CoV-2 infection and the sample date of any positive SARS-CoV-2 PCR result. For analysis, “evidence of previous infection” was defined as individuals reporting a positive SARS-Cov-2 PCR test or an N-antibody level ≥0.4 in submitted blood sample(s) (15).

### Data management and analysis

Data were managed in Microsoft Access, analysed using STATA/SE v.14.2 and graphs created in RStudio or STATA. Antibody geometric mean concentrations (GMC) were calculated for each group. Within-individual geometric mean ratios (GMR) of antibody responses between the 30-day and 12-week timepoints were calculated using mixed-effects regression on log responses, including a random effect for each individual.

Calculation of antibody GMR between vaccine groups with 95% confidence intervals (CI) allowed non-inferiority to be assessed. Adjustment for covariates (age, sex, schedule) was performed as part of a multivariable regression model on log transformed data. The relationship between S-antibody levels and dosing schedule was explored by comparison of fractional polynomial models, and 1/(days between doses)^2^ was found to be optimal. Interactions between vaccine group and covariates were explored and likelihood ratio tests did not indicate that these interaction terms should be included in the adjusted model.

## Results

### Recruitment, retention, sample return and self-sampling success rate

During March – October 2021, 75,827 individuals were identified in NIMS as receiving heterologous vaccination, 9489 were invited to participate by completing the online survey, 1836 responses were received (response rate, 19.3%) and 656 met the eligibility criteria and were recruited for the evaluation (Figure 1). Among ChAd-BNT recipients, sufficient serum within the specified time frame was available for 216 participants at day 30 and 195 at week 12 after the second vaccine dose. In the BNT-ChAd cohort, the numbers were 110 and 77 respectively. Of those using the self-sampling device, the volume of blood was insufficient for testing in approximately 1 in 5 participants, with older, male participants less likely to return a sufficient blood sample.

**Figure 1.**
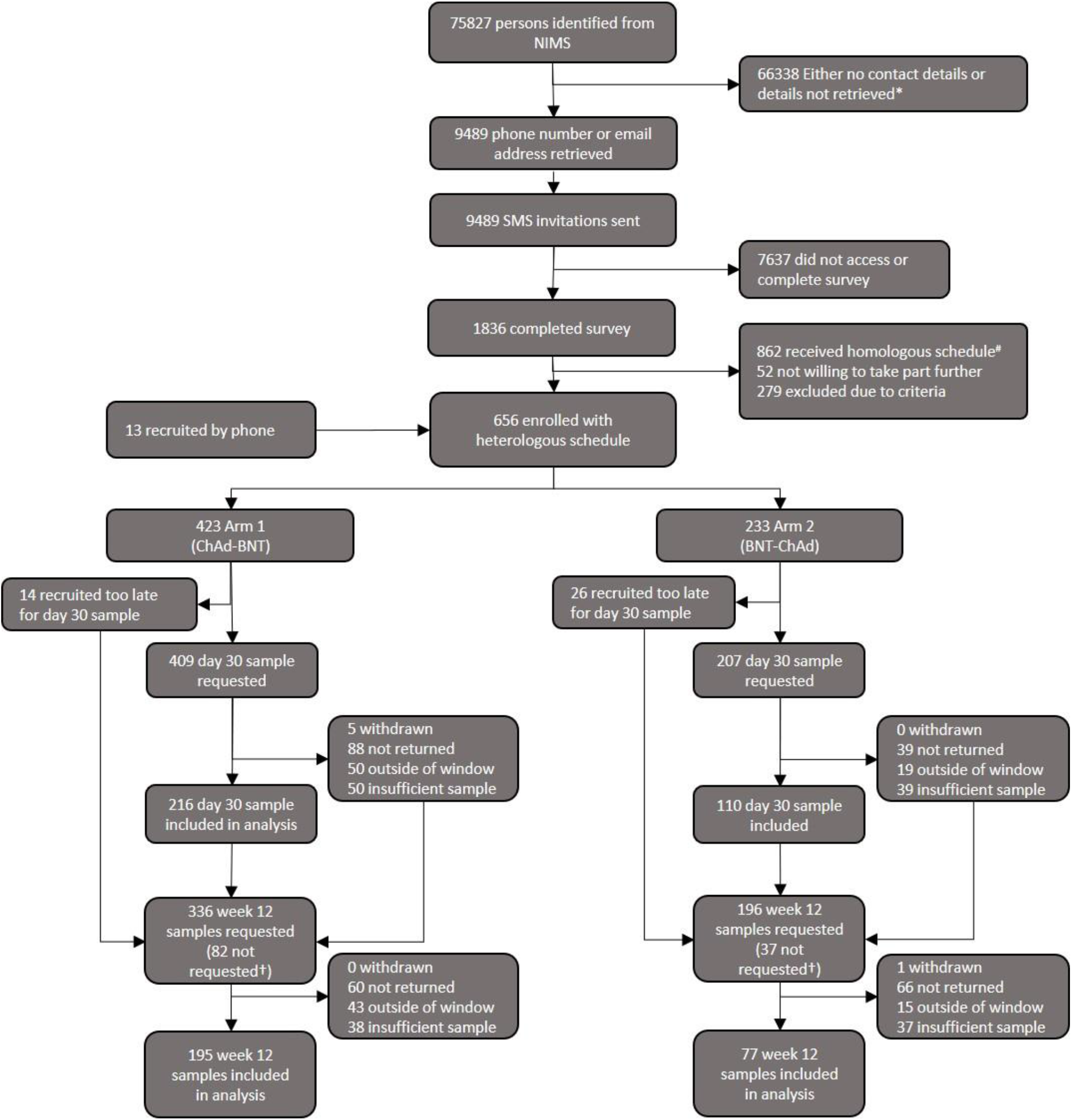
Participant identification, invitation, recruitment, sampling and retention. *Retrieval of contact details were prioritised according to study participant numbers; once one study arm was fully recruited, only contact details of those who were eligible for the other arm were retrieved. ^#^A number of individuals who had received homologous vaccine schedules were invited to complete the same online survey to form a comparator group for a reactogenicity study (6). †Not all participants were invited to provide a sample at week 12 as at the time of writing not all had reached the required length of time following second vaccine dose, or enough samples had already been received for the study group at week 12.

### Demographics of study participants

Participant demographics are summarised in Table 1. The median interval between doses for all participants was 75 days. Individuals receiving a heterologous schedule were younger and with a higher proportion of females than those receiving homologous schedules. Ethnic diversity was highest in the group of participants who received two doses of ChAd, and similar amongst the other study groups.

**Table 1:** Demographic information for individuals with no evidence of natural infection with SARS-CoV-2 for whom at least one anti-S antibody result was available.

| Vaccine schedule | N | Median age, years (IQR) | Sex (if stated) n male (%) | Ethnicity n White (%)<br>n Asian (%)<br>n Black (%)<br>n other (%) | Median time between doses, days (IQR) | n with Ab result at day 30 | n with Ab result at week 12 | n with results at both timepoints |
| --- | --- | --- | --- | --- | --- | --- | --- | --- |
| **No evidence of past infection** |  |  |  |  |  |  |  |  |
| ChAd-ChAd | 121 | 65<br>(54 - 69) | 59<br>(49.2) | 95 (79.2)<br>11 (9.2)<br>9 (7.5)<br>6 (5.0) | 70<br>(54 - 77) | 104 | 44 | 27 |
| BNT-BNT | 135 | 71<br>(69 - 72) | 64<br>(47.4) | 123 (92.5)<br>2 (1.5)<br>1 (0.8)<br>9 (6.7) | 76<br>(70 - 76) | 115 | 49 | 29 |
| ChAd-BNT | 237 | 47<br>(37 - 59) | 55<br>(23.4) | 215 (91.5)<br>7 (3.0)<br>2 (0.9)<br>13 (5.5) | 73<br>(64 - 83) | 191 | 174 | 128 |
| BNT-ChAd | 123 | 51<br>(40 - 63) | 36<br>(29.3) | 112 (93.3)<br>3 (2.5)<br>3 (2.5)<br>5 (4.1) | 79<br>(65 - 99) | 98 | 74 | 49 |
| **Evidence of past infection** |  |  |  |  |  |  |  |  |
| ChAd-ChAd | 32 | 59<br>(53.5 - 66) | 14<br>(43.8) | 21 (65.6)<br>5 (15.6)<br>6 (18.8)<br>0 (0.0) | 72<br>(54 - 77) | 27 | 11 | 6 |
| BNT-BNT | 18 | 71<br>(70 - 72) | 7<br>(38.9) | 13 (72.2)<br>2 (11.1)<br>1 (5.6)<br>2 (11.1) | 76<br>(76 - 77) | 16 | 5 | 3 |
| ChAd-BNT | 33 | 42<br>(31 - 54) | 7<br>(21.2) | 28 (87.5)<br>4 (12.5)<br>0 (0.0)<br>1 (3.0) | 75<br>(59 - 84) | 25 | 21 | 13 |
| BNT-ChAd | 13 | 43<br>(36 - 49) | 5<br>(38.5) | 12 (92.3)<br>1 (7.7)<br>0 (0.0)<br>0 (0.0) | 84<br>(68 - 103) | 12 | 3 | 2 |

Ethnicity was self-described and categories grouped for presentation as follows; White included "White (English/Welsh/Scottish/Northern Irish/British)", "White (Irish)" and "Any other white background". Asian included "Asian/Asian British (Bangladeshi)", "Asian/Asian British (Chinese)", "Asian/Asian British (Indian)" and "Any other Asian background". Black included "Black/African/Caribbean/Black British (African)" and "Black/African/Caribbean/Black British (Caribbean)". Other included "Any other ethnic group", "Any other Mixed/Multiple ethnic background", "Mixed/Multiple ethnic groups (White and Asian)", "Other ethnic group (Arab)" and "prefer not to say".

### Spike Antibody responses

#### Previously Uninfected Individuals

At 30 days after the second vaccine dose, S-antibody GMC (adjusted for schedule, age and sex in a multivariable regression model) were lowest among ChAd-ChAd recipients compared to the other three schedules which included at least one BNT dose (Table 3). Compared to ChAd-ChAd recipients, those receiving ChAd-BNT (GMR 6.29 [95%CI, 5.04-7.85]) and BNT-ChAd (GMR 4.55 [3.56 - 5.81]) had significantly higher S-antibody levels as did those who received BNT-BNT (GMR 5.66 [4.49 - 7.15]), all p<0.001 (Table 3). S-Antibody levels among BNT-BNT recipients were not significantly different compared to ChAd-BNT or BNT-ChAd (Table 3). Comparing between the heterologous vaccine groups, those receiving ChAd-BNT had significantly higher S-antibody levels compared to those receiving BNT-ChAd (GMR 1.38 [95%CI, 1.12 – 1.71]; P=0.003).

**Table 3:** Comparison of adjusted anti-S protein antibody level by vaccine schedule, grouped according to history of SARS-CoV-2 infection.

| Vaccine schedule | Adjusted geometric mean ratio at 30 days (95% CI) | p | Adjusted* geometric mean ratio at 12 weeks (95% CI) | p |
| --- | --- | --- | --- | --- |
| **No evidence of past infection\*** |  |  |  |  |
| **ChAd-ChAd as reference category** |  |  |  |  |
| ChAd-ChAd | 1 (ref) |  | 1 (ref) |  |
| BNT-BNT | 5.66 (4.49 - 7.15) | <0.001 | 6.06 (4.32 - 8.50) | <0.001 |
| ChAd-BNT | 6.29 (5.04 - 7.85) | <0.001 | 5.12 (3.79 - 6.92) | <0.001 |
| BNT-ChAd | 4.55 (3.56 - 5.81) | <0.001 | 4.1 (2.96 - 5.69) | <0.001 |
| **BNT-BNT as reference category** |  |  |  |  |
| ChAd-ChAd | 0.18 (0.14 - 0.22) | <0.001 | 0.16 (0.12 - 0.23) | <0.001 |
| BNT-BNT | 1 (ref) |  | 1 (ref) |  |
| ChAd-BNT | 1.11 (0.88 - 1.40) | 0.376 | 0.85 (0.63 - 1.14) | 0.265 |
| BNT-ChAd | 0.80 (0.62 - 1.03) | 0.087 | 0.68 (0.49 - 0.93) | 0.016 |
| **ChAd-BNT as reference category** |  |  |  |  |
| ChAd-ChAd | 0.16 (0.13 - 0.20) | <0.001 | 0.20 (0.14 - 0.26) | <0.001 |
| BNT-BNT | 0.90 (0.71 - 1.14) | 0.376 | 1.18 (0.88 - 1.59) | 0.265 |
| ChAd-BNT | 1 (ref) |  | 1 (ref) |  |
| BNT-ChAd | 0.72 (0.59 - 0.89) | 0.003 | 0.80 (0.64 - 1.00) | 0.053 |
| **BNT-ChAd as reference category** |  |  |  |  |
| ChAd-ChAd | 0.22 (0.17 - 0.28) | <0.001 | 0.24 (0.18 - 0.34) | <0.001 |
| BNT-BNT | 1.24 (0.97 - 1.60) | 0.087 | 1.48 (1.07 - 2.03) | 0.016 |
| ChAd-BNT | 1.38 (1.12 - 1.71) | 0.003 | 1.25 (1.00 - 1.56) | 0.053 |
| BNT-ChAd | 1 (ref) |  | 1 (ref) |  |
| **Evidence of past infection#** |  |  |  |  |
| **ChAd-ChAd as reference category** |  |  |  |  |
| ChAd-ChAd | 1 (ref) |  |  |  |
| BNT-BNT | 2.23 (1.26 - 3.95) | 0.006 |  |  |
| ChAd-BNT | 1.26 (0.70 - 2.26) | 0.439 |  |  |
| BNT-ChAd | 0.69 (0.36 - 1.33) | 0.265 |  |  |
| **BNT-BNT as reference category** |  |  |  |  |
| ChAd-ChAd | 0.45 (0.25 - 0.79) | 0.006 |  |  |
| BNT-BNT | 1 (ref) |  |  |  |
| ChAd-BNT | 0.56 (0.28 - 1.15) | 0.116 |  |  |
| BNT-ChAd | 0.31 (0.14 - 0.67) | 0.003 |  |  |
| **ChAd-BNT as reference category** |  |  |  |  |
| ChAd-ChAd | 0.79 (0.45 - 1.38) | 0.405 |  |  |
| BNT-BNT | 1.72 (0.86 - 3.44) | 0.123 |  |  |
| ChAd-BNT | 1 (ref) |  |  |  |
| BNT-ChAd | 0.45 (0.24 - 0.82) | 0.010 |  |  |
| **BNT-ChAd as reference category** |  |  |  |  |
| ChAd-ChAd | 1.77 (0.93 - 3.36) | 0.083 |  |  |
| BNT-BNT | 3.87 (1.83 - 8.19) | <0.001 |  |  |
| ChAd-BNT | 2.25 (1.22 - 4.14) | 0.01 |  |  |
| BNT-ChAd | 1 (ref) |  |  |  |

\*Adjusted for age group, sex and vaccine schedule.
#Adjusted for age group and sex. Adjustment for dosing schedule was checked and dropped.

S-antibody GMC at 12 weeks were around 50% lower than at 30 days after the second vaccine dose for all four schedules, consistent with waning of circulating antibodies (Table 2). Notably, antibody levels at 12 weeks post-dose two were significantly higher when the schedule contained at least one BNT dose compared to ChAd-ChAd, and those receiving BNT as the second dose had significantly higher antibody levels than those receiving ChAd as the second dose (Table 3). At this timepoint, S-antibody levels were lower for BNT-ChAd (GMR 0.68 [0.49-0.93]; P=0.016) compared to BNT-BNT. There was no difference in S-antibody levels at either time point between BNT-BNT and ChAd-BNT. (Table 3, Figure 2).

**Table 2:** Geometric mean anti-S antibody levels and within-individual geometric mean ratio of responses relative to 30 days (±9 days) post dose 2 of vaccine (results omitted if <5 samples per group).

| Vaccine schedule | Study time point, post dose 2 (range) | N | Geometric mean Anti-S antibody level (95% CI) | Within-individual geometric mean ratio of response relative to 30 days (95% CI) |
| --- | --- | --- | --- | --- |
| **No evidence of past infection** |  |  |  |  |
| **ChAd-ChAd** | 30 days (21-39 days) | 104 | 862<br>(694 - 1069) | 1 (ref) |
|  | 12 weeks (10-14 weeks) | 44 | 360<br>(259 - 501) | 0.53<br>(0.45 - 0.61) |
| **BNT-BNT** | 30 days (21-39 days) | 115 | 5377<br>(4596 - 6289) | 1 (ref) |
|  | 12 weeks (10-14 weeks) | 49 | 2549<br>(2036 - 3192) | 0.5<br>(0.46 - 0.54) |
| **ChAd-BNT** | 30 days (21-39 days) | 191 | 6233<br>(5522 - 7035) | 1 (ref) |
|  | 12 weeks (10-14 weeks) | 174 | 2396<br>(2102 - 2732) | 0.39<br>(0.36 - 0.41) |
| **BNT-ChAd** | 30 days (21-39 days) | 98 | 4776<br>(4066 - 5610) | 1 (ref) |
|  | 12 weeks (10-14 weeks) | 74 | 1930<br>(1617 - 2303) | 0.45<br>(0.41 - 0.48) |
| **Evidence of past infection** |  |  |  |  |
| **ChAd-ChAd** | 30 days (21-39 days) | 27 | 10320<br>(7171 - 14853) | 1<br>(ref) |
|  | 12 weeks (10-14 weeks) | 11 | 5271<br>(2902 - 9574) | 0.62<br>(0.5 - 0.76) |
| **BNT-BNT** | 30 days (21-39 days) | 16 | 20983<br>(13756 - 32008) | 1 (ref) |
|  | 12 weeks (10-14 weeks) | 5 | 6210<br>(2636 - 14631) | 0.55<br>(0.48 - 0.63) |
| **ChAd-BNT** | 30 days (21-39 days) | 25 | 16349<br>(11434 - 23377) | 1 (ref) |
|  | 12 weeks (10-14 weeks) | 21 | 7266<br>(4341 - 12162) | 0.49<br>(0.42 - 0.57) |
| **BNT-ChAd** | 30 days (21-39 days) | 12 | 8553<br>(4807 - 15219) | - |
|  | 12 weeks (10-14 weeks) | 3 | - | - |

Note: All samples were anti-S antibody positive at all time points.

**Figure 2.**
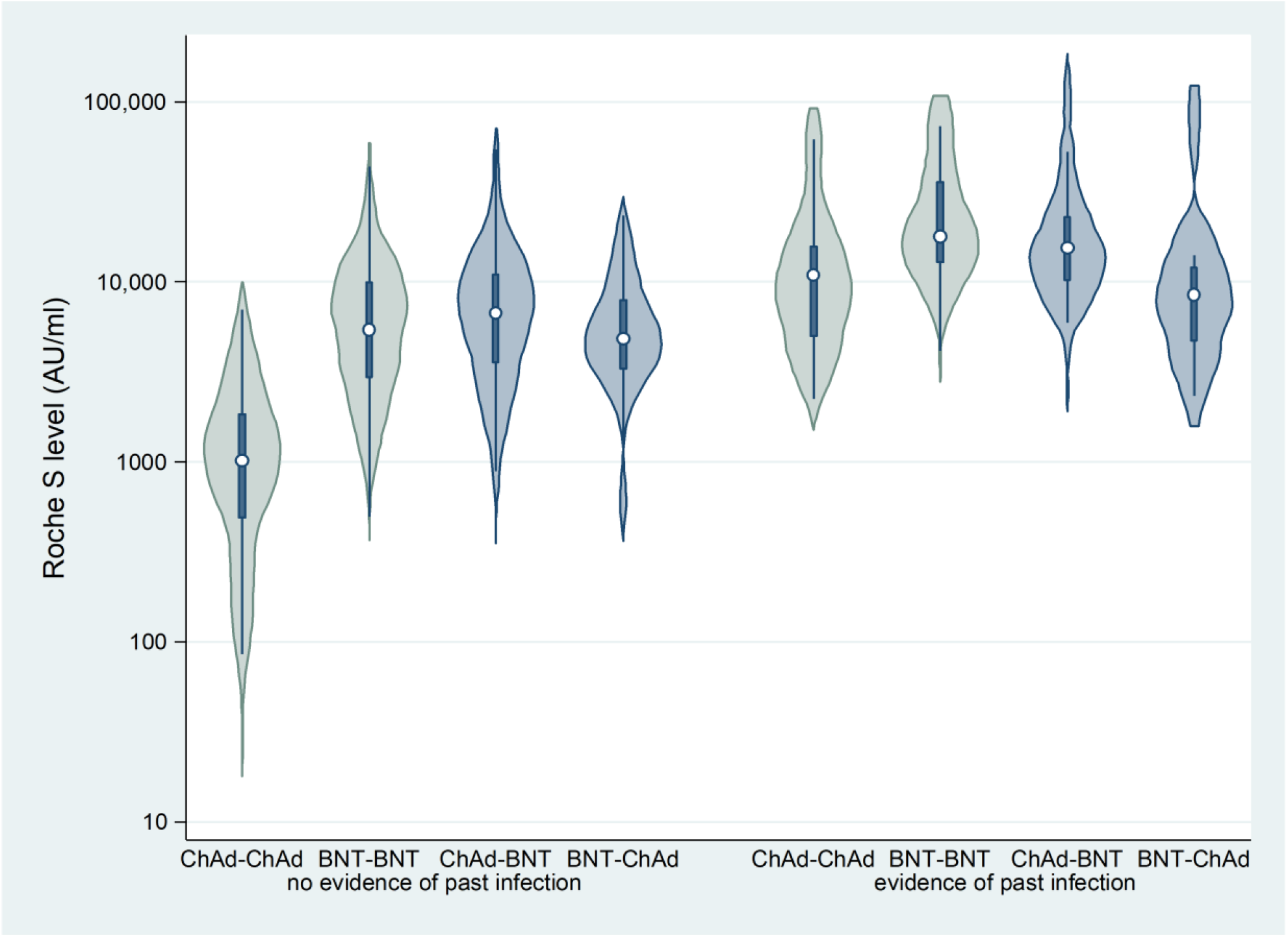
Violin plots depicting the distribution of anti-S antibody levels at 30 days (±9 days) post dose 2 of vaccine. (White dots represent the median, dark blue bars show the IQR).

#### Previously Infected Individuals

Unlike infection-naïve adults, age and sex were not significantly associated with antibody levels at 30 days after the second vaccine dose in previously infected individuals. The model was not adjusted for dosing schedule, as inclusion of this variable did not improve the model fit.

In previously infected adults, S-antibody levels were higher than those observed in the infection-naïve group at all time-points and with all vaccine schedules.

At 30 days after dose 2, S-antibody levels compared to ChAd-ChAd recipients were only significantly higher for BNT-BNT recipients (2.23 [1.26-3.95] P=0.006; Table 3) but not ChAd-BNT or BNT-ChAd. Additionally, among heterologous schedule recipients, S-antibody levels were significantly higher in those receiving ChAd-BNT compared to BNT-ChAd (GMR, 2.25; [1.22-4.14]; P=0.010). As observed with infection naïve individuals, antibody responses were higher in previously infected individuals who received BNT as their second dose.

## Discussion

We prospectively recruited adults who had received heterologous COVID-19 vaccine schedules as part of the UK national vaccine rollout which began in December 2020. Both BNT and ChAd were available initially and, whilst national recommendations were to use the same vaccine product for both doses, some adults received a heterologous schedule, primarily following a serious adverse reaction to their first dose. The emergence of rare reports of VITT after ChAd vaccination prompted an urgent assessment of the reactogenicity and immunogenicity of heterologous extended schedules. In more recent months, reports of myocarditis following mRNA vaccines further highlights the importance of evaluating different combinations of mixed schedules. Our real-world data demonstrate that two BNT doses provide the highest antibody responses at 30 days after two doses given according to the UK-recommended extended schedule, while two ChAd doses elicit the lowest circulating antibody level. Heterologous schedules induced similar antibody responses as two BNT doses, although antibody levels were significantly higher after Chad-BNT compared to BNT-ChAd at least up to 12 weeks after the second vaccine dose. Finally, previously infected individuals had very high antibody responses irrespective of the immunisation schedule, and these higher antibody levels were maintained for at least 12 weeks after the second vaccine dose.

With the global pandemic still on-going, access to COVID-19 vaccines remains restricted especially in lower- and middle-income countries. ChAd was developed specifically to provide an affordable vaccine globally and has the added advantage of not requiring ultralow storage temperatures. With concerns about the rare but severe VITT associated with ChAd, a lot of attention has been focused on heterologous immunisation to enable adults who had received a single ChAd dose to complete their immunisation with an alternative vaccine. Our findings add to the growing body of evidence describing very high antibody responses after mRNA vaccination following a single dose of ChAd; far greater than ChAd-ChAd, and only marginally lower BNT-BNT (16).

The immunological correlates of protection against infection with SARS-CoV-2 or severe disease outcomes have not yet been determined and, therefore, increased antibody levels above a certain threshold may not necessarily translate into clinically relevant benefit. Importantly, in our cohort all participants receiving any vaccine combination had high S-antibodies at all timepoints after vaccination. High antibody neutralising activity has also been reported after two vaccine doses even with heterologous vaccine schedules and, similar to mRNA vaccines, the homologous ChAd-ChAd schedule also induces robust cellular responses (16) (10). Cellular immunity likely contributes substantially towards the real-world vaccine effectiveness of ChAd which, despite the lower post-vaccination circulating antibody levels, remained 44% effective against symptomatic disease compared to 59.5% for BNT at >140 days after two doses, using the UK extended 8-12 week interval schedule (17). Reassuring, too, is that both the homologous ChAd and homologous BNT schedules retain high vaccine effectiveness against more severe outcomes of infection such as hospitalisation and death, even with the more transmissible and more virulent delta variant (18).

Our evaluation is different to most of the currently published studies in that we evaluated both heterologous schedules contemporaneously with the two corresponding homologous schedules, and augments the findings of the only other study (COMCOV) reporting immune responses at 28 days post-dose 2 using the four different schedules by including extended antibody follow-up data up to 12 weeks after vaccination as well as antibody responses in previously-infected participants (10). Additionally, unlike the COMCOV study our cohort is unique in that the participants had an extended 12-week interval between doses, which has shown to provide higher boosting and longer protection compared to the shorter 3-4 weeks interval authorised for mRNA vaccines (11) (19), and represents the current recommended schedule in the UK. Not only were we able to confirm increased post-vaccination antibody levels in adults receiving ChAd-ChAd, ChAd-BNT and BNT-BNT, but we also showed significant differences in antibody responses between the two heterologous schedules, such that ChAd-BNT recipients achieved higher S-antibody levels than BNT-ChAd recipients. Interestingly, in the COMCOV study, BNT-ChAd recipients had the greatest expansion of vaccine-induced antigen-specific T cells in the peripheral circulation at 28 days after the second vaccine dose which, compared to other ChAd combinations, may result in equivalent or better protection against the virus as circulating antibody levels decline with time since vaccination.

Our data suggest a steeper decline in spike-antibodies in the heterologous schedule groups compared to their homologous counterparts (i.e. ChAd-BNT vs ChAd-ChAd and BNT-ChAd vs BNT-BNT), which may indicate reduced longevity of protection. However, this should be interpreted with caution as the sampling strategy of the study was not designed to assess kinetics of the humoral response. The time-points evaluated were selected with a view that the initial recall antibody expansion would have happened by day 30, with a contraction phase occurring between day 30 and week 12. BNT and ChAd are expected to have different kinetics, in terms of the primary and recall immune responses generated, owing to their differing modes of action (20). It is not yet known at which point in time the contraction of the recall response after either vaccine is complete, nor is it certain that a lower level of circulating antibody results in a less effective recall immune response if/when challenged with infection.

A recent Swedish population-based surveillance reported 67% vaccine effectiveness against symptomatic disease for ChAd-BNT and 79% for ChAd-mRNA-1273 (Spikevax, Moderna), compared to 50% for ChAd-ChAd (21). Based on our immunogenicity data, both heterologous schedules are likely to provide higher protection against COVID-19 than ChAd-ChAd. Recommendations for a heterologous schedule, however, have to be balanced with the higher reactogenicity rates after the second vaccine dose reported by us and others (6) (7). Additionally, the low but severe risk of VITT after the first ChAd dose in young adults must be considered, along with the potential recurrence of myocarditis with the second mRNA dose in young adults who experienced this rare adverse event after their first mRNA vaccine dose. Both scenarios exemplify the utility of the option for heterologous vaccine schedules in specific circumstances.

### Strengths and Limitations

The strength of this evaluation is the use of national surveillance data to rapidly identify and recruit adults who had received a heterologous vaccine schedule as part of the national COVID-19 immunisation programme. This real-world approach was effective and efficient, facilitating speedy identification, enrolment, and timely sample collection after the second vaccine dose. The use of self-sampling blood collection devices removed the need for phlebotomy and allowed participation of individuals from across the country, increasing the generalisability of our findings to the UK population. The volume of blood collected by self-sampling was sufficient for serological assays but not for additional studies such as cellular immune responses.

There are some limitations. The recruited participants may not be representative of the general population since they deviated from the national recommendation to receive the same vaccine product for both doses, usually because of a severe reaction after the first dose. Also, as this was not a clinical trial, we were unable to collect blood samples prior to the second vaccine dose and blood sampling times were more variable, with insufficient sample volume in up to 20% of returned self-sampling devices. Finally, the infection status of the participants was not known at recruitment, leading to the small sample size for previously infected individuals, especially when sub-grouped by vaccine schedule.

### Implications and Conclusions

These real-world findings provide additional reassurance of high antibody responses after a heterologous schedule, allowing increased flexibility for programme delivery especially in the context of global supply constraints. A heterologous schedule provides higher antibody levels than ChAd-ChAd, with higher antibody responses after ChAd-BNT compared to BNT-ChAd. Real-world data also confirm higher vaccine effectiveness against symptomatic disease with ChAd-BNT and ChAd-mRNA-1273 compared to ChAd-ChAd, which is salient information for countries that avoided giving a second ChAd dose because of safety concerns relating to VITT, instead offering an mRNA vaccine as the second dose (5) (7) (8) (9). We additionally demonstrate superior antibody responses following BNT-ChAd compared to ChAd-ChAd, with significant difference maintained for at least 12 weeks post-vaccination. Taken together, our findings and those of others support the use of heterologous schedules in national immunisation programmes, especially in countries with limited vaccine supply. Notably, the United States FDA recently approved heterologous prime-boost vaccination schedules (22). Our findings also support the recent UK decision to offer an mRNA vaccine as a third booster dose to adults at least 3 months after a two-dose primary schedule (4). We recently reported very high antibody boosting in adults receiving either ChAd-ChAd or BNT-BNT primary vaccinations, reaching similar levels between the two groups (23). These findings are consistent with the high vaccine effectiveness achieved by the booster dose against symptomatic disease in the UK (17), and elsewhere (24). Response to booster vaccination following heterologous primary immunisation schedules remain unknown. The participants of this evaluation will continue to be followed up after receiving their booster as part of the UK national immunisation programme.

## Data Availability

All data produced in the present study are available upon reasonable request to the authors

## Acknowledgements

The authors would like to thank all study participants. The authors are also grateful to UKHSA staff who helped with the study: Adolphe Bukasa, Julie Brough, Paul Charter, Deborah Cohen, Teresa Gibbs, Natalie Mensah, Silvina Omisore, Charlotte Ryan and Molly Viggars.

## Funding

This surveillance was internally funded by Public Health England (now UK Health Security Agency) and did not receive any specific grant funding from agencies in the public, commercial or not-for-profit sectors.

## Conflict of interest

SW has previously (2009 – 2012) worked on a non-vaccine related clinical study funded by Pfizer Global via an academic institution; the subject area was outside of the submitted work.

## Authors’ contributions

GA, SL, KB and MR contributed to the design of the research, SW, AP, LP, CW, HC, RS and LW contributed to implementation of the study, SW, HW, GA and KB analysed the data and wrote the manuscript with input from all authors.

## Notes

### Competing Interest Statement

SW has previously (2009 to 2012) worked on a non-vaccine related clinical study funded by Pfizer Global via an academic institution; the subject area was outside of the submitted work.

### Author Declarations

The study protocol was subject to an internal review by the Public Health England Research Ethics and Governance Group and was found to be fully compliant with all regulatory requirements (study number NR0227). As no regulatory issues were identified, and ethical review is not a requirement for this type of work, it was decided that a full ethical review would not be necessary. Public Health England has legal permission, provided by Regulation 3 of The Health Service (Control of Patient Information) Regulations 2002, to process patient confidential information for national surveillance of communicable diseases and as such, individual patient consent is not required to access records. All necessary patient/participant consent has been obtained by those who completed the questionnaire and provided blood samples, and the appropriate institutional forms have been archived.

